# Impact of Multimodal Prompt Elements on Diagnostic Performance of GPT-4(V) in Challenging Brain MRI Cases

**DOI:** 10.1101/2024.03.05.24303767

**Authors:** Severin Schramm, Silas Preis, Marie-Christin Metz, Kirsten Jung, Benita Schmitz-Koep, Claus Zimmer, Benedikt Wiestler, Dennis M. Hedderich, Su Hwan Kim

## Abstract

**Background:** Recent studies have explored the application of multimodal large language models (LLMs) in radiological differential diagnosis. Yet, how different multimodal input combinations affect diagnostic performance is not well understood.

**Purpose:** To evaluate the impact of varying multimodal input elements on the accuracy of GPT-4(V)-based brain MRI differential diagnosis.

**Methods:** Thirty brain MRI cases with a challenging yet verified diagnosis were selected. Seven prompt groups with variations of four input elements (image, image annotation, medical history, image description) were defined. For each MRI case and prompt group, three identical queries were performed using an LLM-based search engine (© PerplexityAI, powered by GPT-4(V)). Accuracy of LLM-generated differential diagnoses was rated using a binary and a numeric scoring system and analyzed using a chi-square test and a Kruskal-Wallis test. Results were corrected for false discovery rate employing the Benjamini-Hochberg procedure. Regression analyses were performed to determine the contribution of each individual input element to diagnostic performance.

**Results:** The prompt group containing an annotated image, medical history, and image description as input exhibited the highest diagnostic accuracy (67.8% correct responses). Significant differences were observed between prompt groups, especially between groups that contained the image description among their inputs, and those that did not. Regression analyses confirmed a large positive effect of the image description on diagnostic accuracy (p ≪ 0.001), as well as a moderate positive effect of the medical history (p < 0.001). The presence of unannotated or annotated images had only minor or insignificant effects on diagnostic accuracy.

**Conclusion:** The textual description of radiological image findings was identified as the strongest contributor to performance of GPT-4(V) in brain MRI differential diagnosis, followed by the medical history. The unannotated or annotated image alone yielded very low diagnostic performance. These findings offer guidance on the effective utilization of multimodal LLMs in clinical practice.

## Introduction

Rapid developments in the field of large language models (LLMs) have attracted significant interest in potential medical applications. In radiology, LLMs have been explored for the generation of radiology reports (1–3), differential diagnosis based on case presentations (4–7), the automated definition of imaging protocols (8,9), the extraction of structured data from radiology reports (10,11) and more.

Recent studies have further explored the diagnostic application of multimodal LLMs (also called ‘vision-language models’) that are able to ingest not only text but also image data as input (12–20). However, several studies demonstrated low performance of *Generative Pre-trained Transformer 4 Vision* (GPT-4V) by OpenAI in differential diagnosis based on various types of radiological images (12,16,18,20,21). These studies evaluated queries with only images (12,18,20,21) or clinical information and images (16) as input.

Importantly, the provided types of model input were shown to have a relevant impact on diagnostic performance. One study showed higher diagnostic performance with multimodal input (image and medical history) as compared to text-only or image-only prompts (17). However, more granular variations in input elements such as textual descriptions of image findings and image annotations have not been investigated systematically.

Therefore, the aim of this study was to evaluate the impact of multimodal input elements on the accuracy of GPT-4(V)-based brain MRI differential diagnosis.

## Methods

### Study Design

Ethical approval was waived by the ethics committee of the Technical University of Munich. Seven different prompt groups with varying input types were defined (Table 1). For each clinical case, up to four representative slices were selected by two neuroradiology residents (SHK and SS) from up to two MRI sequences adequate for each respective pathology and uploaded as model input (groups 1-5). In group 2 and 5, key findings were additionally annotated using up to four arrows. The medical history was obtained from the patient record to simulate a realistic clinical scenario. Descriptions of key image findings were created by SHK and SS and included details about location, morphology, signal behavior, and contrast enhancement in a standardized manner.

**Table 1:** Prompt groups.

| Group No. | Abbreviation | Description |
| --- | --- | --- |
| 1 | I | Image |
| 2 | I + A | Image + Annotation |
| 3 | I + H | Image + Medical History |
| 4 | I + D | Image + Image Description |
| 5 | I + A + H + D | Image + Annotation + Medical History + Image Description |
| 6 | H + D | Medical History + Image Description |
| 7 | D | Image Description |

**Table 2:** Pairwise testing for inter-group differences in binary scores. Values indicate p-values for the respective group differences. All p-values were adjusted for a false-discovery rate of 0.05.

| Chi-Square Test | 1 (I) | 2 (I+A) | 3 (I+H) | 4 (I+D) | 5 (I+A+H+D) | 6 (H+D) |
| --- | --- | --- | --- | --- | --- | --- |
| 2 (I+A) | 1.000 |  |  |  |  |  |
| 3 (I+H) | 0.162 | 0.162 |  |  |  |  |
| 4 (I+D) | << 0.001 | << 0.001 | 0.004 |  |  |  |
| 5 (I+A+H+D) | << 0.001 | << 0.001 | 0.002 | 1.000 |  |  |
| 6 (H+D) | << 0.001 | << 0.001 | 0.007 | 1.000 | 1.000 |  |
| 7 (D) | << 0.001 | << 0.001 | 0.007 | 1.000 | 1.000 | 1.000 |

### Case Selection

A total of 30 brain MRI cases with challenging yet definitive diagnoses were selected from the local imaging database. Diagnoses had been verified either by histopathology or through the independent agreement of at least two senior neuroradiologists. Scans had been obtained between 01/01/2016 and 12/31/2023. 20 out of 30 cases have been published previously (20). An overview of clinical cases with respective medical histories and image descriptions is provided in Supplement 1.

### Queries and Prompt Design

PerplexityAI (© Perplexity AI Inc., San Francisco, USA), served as the query interface. Search queries were processed by GPT-4(V), OpenAI’s latest multimodal LLM. The base prompt was formulated as follows: “*You are a senior neuroradiologist. Below, you will find information regarding a brain MRI scan. Based on this information, identify the three most likely differential diagnoses, ranked by their likelihood. Present your findings in a table format with the following columns: ‘Rank’, ‘Differential Diagnosis’, and ‘Explanation’*.*”* For each MRI case and prompt group, three identical queries were performed to account for probabilistic variations of the LLM output. Before each query, a new thread was started to exclude undesired influences of prior queries on the LLM response.

### Analysis

Two distinct scoring systems were applied to evaluate the accuracy of differential diagnoses. First, LLM responses were classified as “correct” if the correct diagnosis appeared within the top three suggestions, or “incorrect” if it did not (binary scoring system). Second, a score from 0 to 3 was assigned based on the rank of the correct diagnosis within the LLM response (numeric scoring system). Ratings for edge cases where the suggested diagnosis was only partially correct were determined through consensus by SHK and SS (e.g. the response “cerebral microbleeds” was rated as incorrect in a case with diffuse axonal injury).

Statistical analyses were conducted in RStudio (RStudio, 2022.02.3 Build 492; © 2009 -2022 RStudio, PBC). Statistical significance was set at p < 0.05. Binary and numeric scores were reported using descriptive statistics.

To compare scores between prompt groups, two tests were applied on the median scores across the three measurements. For the binary scoring system, a chi-square test was performed over all groups with subsequent pairwise testing. For the numeric scoring system, a Kruskal-Wallis test was conducted across all groups followed by pairwise testing using Dunn’s test (21). For both scoring systems, results were adjusted to a false-discovery rate of 0.05 by employing the Benjamini-Hochberg procedure (22).

Additionally, two mixed-effects regression models (binomial logistic model for binary scores, ordinal cumulative link model for numeric scores) were used to determine the contribution of individual input elements (image [I], annotation [A], medical history [H], and image description [D]) (23). The four input elements were modelled as explanatory variables, while the binary and numeric score were treated as response variables. For each case, a random intercept was included to model heterogeneities between individual cases and to factor in that three identical prompts were used for each case and prompt group.

## Results

### Descriptive Statistics

Prompt group 5 (I + A + H + D) exhibited the highest diagnostic accuracy (67.8% correct responses, median numeric score of 2). The three other groups containing the image description as input followed: group 6 (H + D), group 4 (I + D), and group 7 (D) yielded 61.1%, 60.0%, 56.7% correct responses and a median numeric score of 2, 2, and 1, respectively. Group 3 (I + H) followed with a big margin, with 21.1% correct responses and a median numeric score of 0. Group 1 (I) and group 2 (I + A) revealed the lowest diagnostic accuracy with only 4.4% and 2.2% correct responses, and a median numeric score of 0 for both (Figure 1).

**Figure 1:**
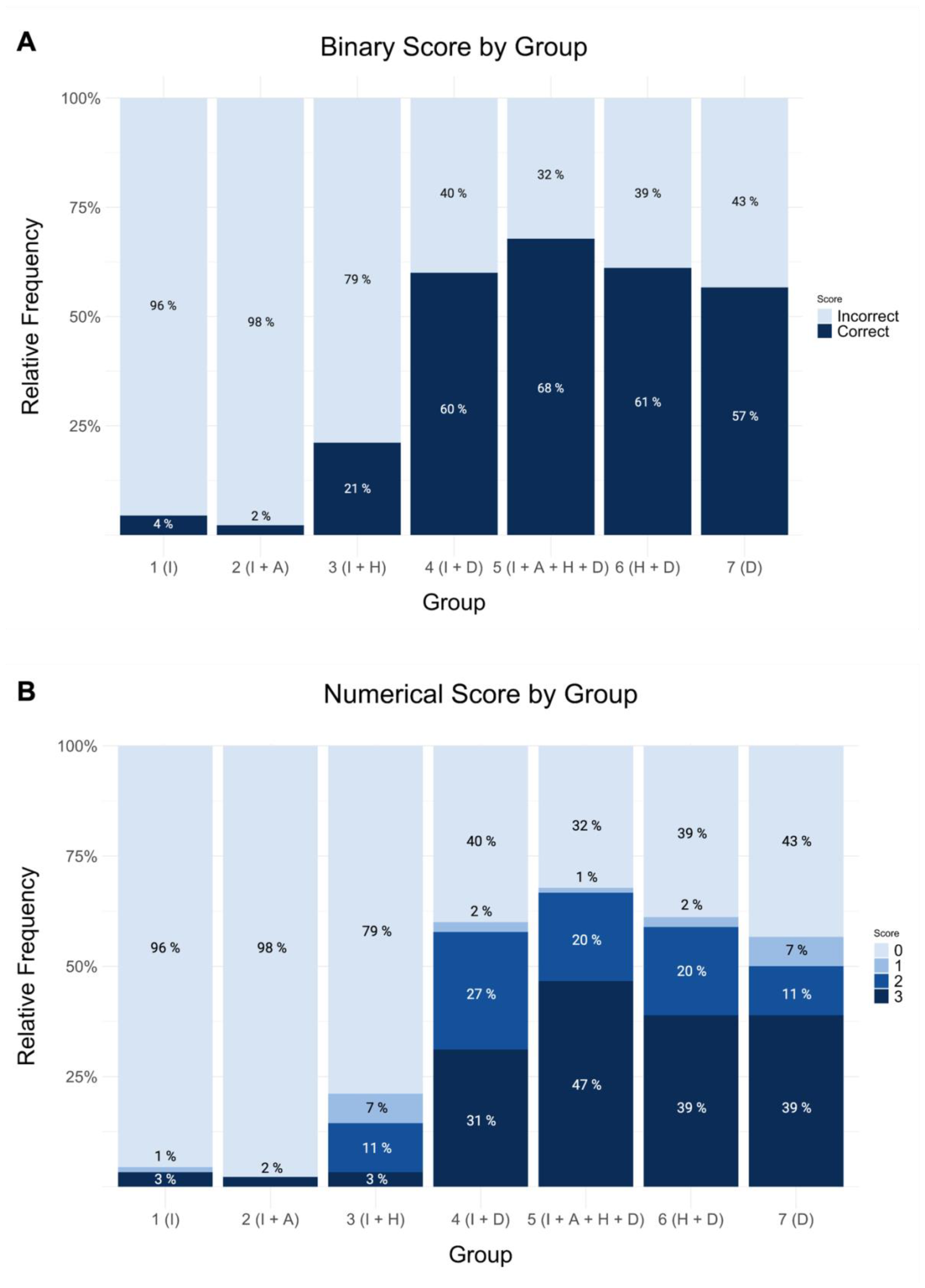
Diagnostic accuracy. A: Binary scores. B: Numeric scores (0-3). 1: Image only. 2: Image + Annotation. 3: Image + Medical History. 4: Image + Image Description. 5: Image + Annotation + Medical History + Image Description. 6: Medical History + Image Description. 7: Image Description.

**Figure 2:**
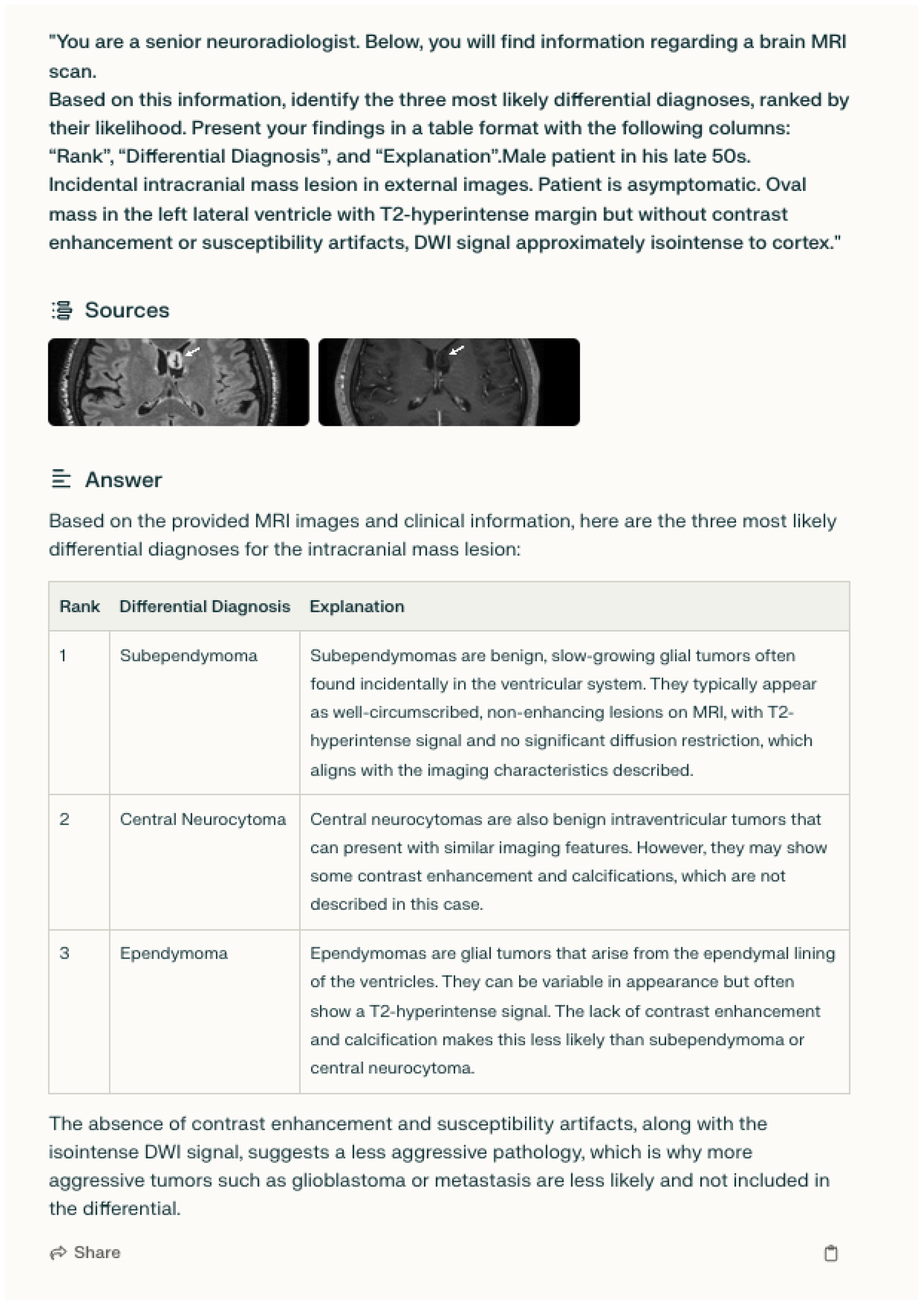
Sample query. The prompt includes the annotated MRI images, the medical history, and an image description (prompt group 5). The correct diagnosis in this case was subependymoma.

### Inferential Statistics

Groups 4-7, all of which contained the image description among their inputs, consistently showed significantly higher diagnostic accuracy over groups 1-3, as measured by median binary (p < 0.01) and numeric score (p < 0.001). No significant differences in median score were observed between groups 1-3.

The binomial logistic mixed-effects regression model was fitted to 630 individual measurements and retained 624 residual degrees of freedom. The model indicated a large positive effect of D (coefficient 4.420; p << 0.001), a moderate positive effect of H (coefficient 1.1960; p < 0.001), a minor positive effect of I (coefficient 0.761; p < 0.05), and a minor negative effect of A (coefficient -0.801; p < 0.05) in reference to the binary score. The cumulative link regression model for numeric scores was similarly fitted to 630 measurements and retained 622 residual degrees of freedom. Similarly to the binomial model, we observed a large positive effect for D (coefficient 4.564; p << 0.001) and a moderate positive effect of H (coefficient 1.042; p < 0.001) on numeric scores. Neither I nor A had a significant effect on the numeric score. Model details are presented in Table 4.

**Table 3:** Pairwise testing for inter-group differences in numerical scores. Values indicate p-values for the respective group differences. All p-values were adjusted for a false-discovery rate of 0.05.

| Dunn's Test | 1 (I) | 2 (I+A) | 3 (I+H) | 4 (I+D) | 5 (I+A+H+D) | 6 (H+D) |
| --- | --- | --- | --- | --- | --- | --- |
| 2 (I+A) | 0.500 |  |  |  |  |  |
| 3 (I+H) | 0.265 | 0.285 |  |  |  |  |
| 4 (I+D) | << 0.001 | << 0.001 | < 0.001 |  |  |  |
| 5 (I+A+H+D) | << 0.001 | << 0.001 | << 0.001 | 0.313 |  |  |
| 6 (H+D) | << 0.001 | << 0.001 | < 0.001 | 0.500 | 0.343 |  |
| 7 (D) | << 0.001 | << 0.001 | < 0.001 | 0.525 | 0.334 | 0.527 |

**Table 4:**
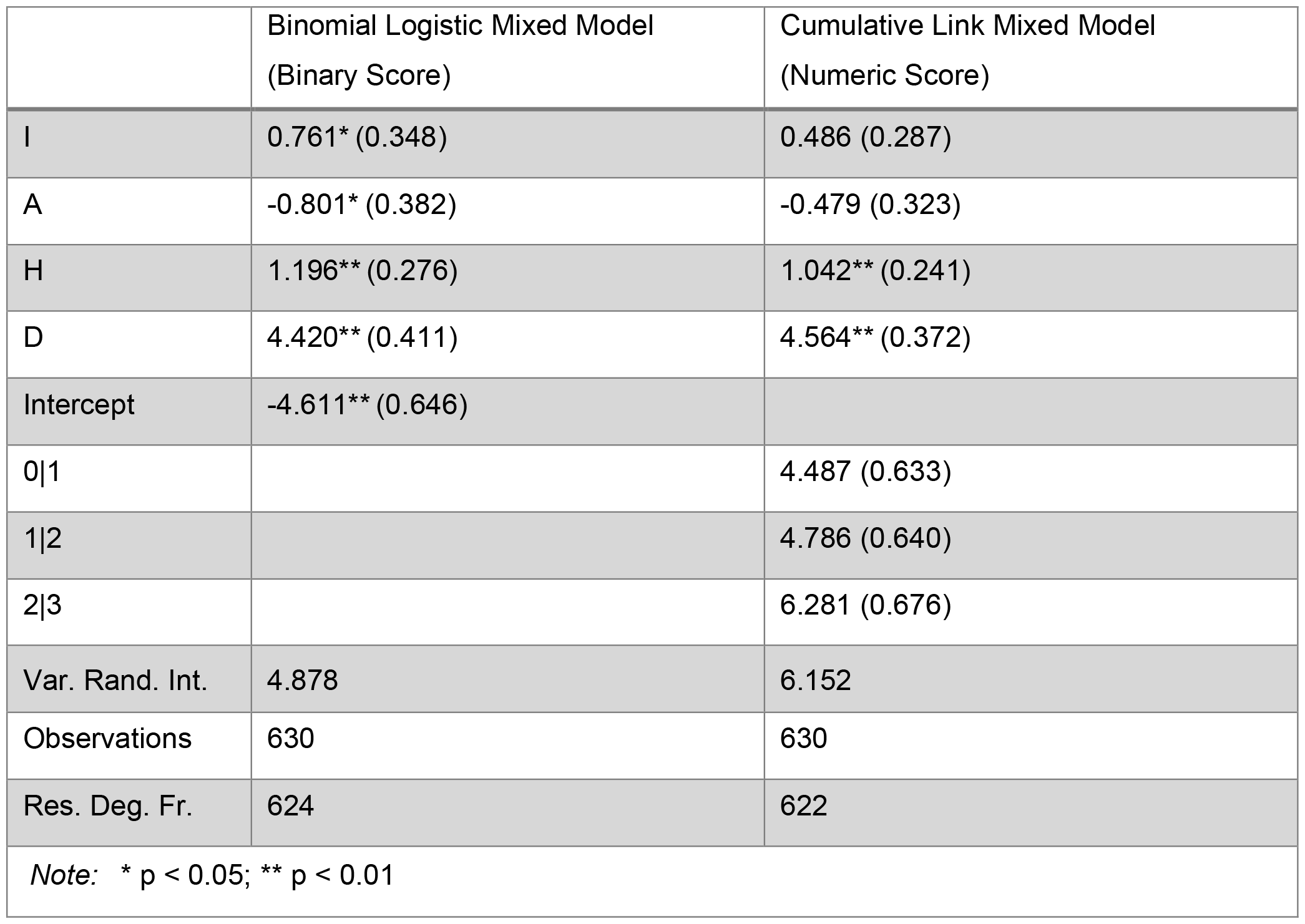
Parameters of the regression models fitted to the binary and numeric scores. Values indicate coefficients and intercepts, with values in brackets indicating the respective standard error. Abbreviations: Annotation (A), Description (D), Medical History (H), Image (I).

## Discussion

This study aimed to investigate the impact of varying input elements on the accuracy of GPT-4(V)-based brain MRI differential diagnosis. In summary, our findings suggest that the image description has the strongest influence on diagnostic performance of GPT-4(V) by far, followed by the medical history. In contrast, diagnostic accuracy was very low with annotated or non-annotated MRI images alone, highlighting the importance of expert image descriptions even in multimodal settings.

This is in line with several prior studies reporting low diagnostic performance of GPT-4(V) with radiological images alone as input (12,18,20,21). Consistent with the findings of Schubert et al, we observed that the combination of medical history and radiological images yielded higher diagnostic accuracy than images alone, although the difference did not reach statistical significance (17).

Remarkably, our study identified the textual description of radiological image findings as the strongest contributor to diagnostic performance, exceeding other input factors by a considerable margin. One possible explanation is that GPT-4(V)’s training data likely contains only a limited quantity of radiological images with high-quality labels, unlike abundant textual content on radiological imaging features of various diagnoses. It is yet to be determined whether utilization of LLM-generated image descriptions as input for sequential LLM queries could increase diagnostic accuracy.

Importantly, interactions between radiologists and multimodal LLMs remain to be investigated. In a previous study, we observed that inaccurate or incomplete LLM prompts phrased by human users can result in misleading LLM responses (22). Future investigations should examine whether systematic education of radiologists on optimal LLM prompting strategies can translate into improved diagnostic accuracy of LLM-enhanced human readers.

### Limitations

This study has several limitations.

First, GPT-4(V) is a generic, non-domain-specific LLM. Multimodal LLMs specifically trained for medicine or radiology are likely to exhibit superior performance in radiological differential diagnosis, although the lack of high-quality data remains a barrier for their development. Second, the brain MRI cases evaluated in this study were highly complex, and the diagnostic performance of GPT-4(V) is presumably higher in an average brain MRI case sample. Third, image descriptions were created in knowledge of the correct diagnosis and might therefore be biased, although terminology highly specific to the true diagnosis had been avoided. The level of variability of image descriptions among radiology readers in a realistic clinical scenario and its impact on LLM-generated differential diagnoses are yet to be examined.

Fourth, despite choosing representative slices in MRI sequences adequate for each respective diagnosis, other slice selections might have led to different results. A full 3D representation of each pathology could potentially increase diagnostic performance. Further studies may consider a dedicated evaluation of this possibility.

Fifth, the regression analyses do not directly reflect the far more complex statistical processes underlying LLM response generation. For example, different wordings of the prompt elements might have led to different results, since each input is processed as a joint input unit. Therefore, the findings should be understood as observations on the purely functional level that do not extend to the underlying algorithmic computations.

In conclusion, this study provides early insights into the role of distinct prompt inputs on the diagnostic performance of LLMs. Our findings inform effective application of LLMs in clinical practice. The presented methodological approach may serve as a benchmark for similar studies evaluating further clinical use cases.

## Data Availability

All data produced in the present study are available upon reasonable request to the authors

## Notes

### Competing Interest Statement

The authors have declared no competing interest.

### Funding Statement

This study did not receive any funding

### Author Declarations

Ethics committee/IRB of the Technical University of Munich waived ethical approval for this work.

